# The Cambridge Questionnaire for Apathy and Impulsivity Traits (CamQUAIT): a novel assessment tool for frontotemporal lobar degeneration-related syndromes

**DOI:** 10.1101/2024.07.01.24309762

**Authors:** CJ Lansdall, R Williams, I Coyle-Gilchrist, AG Murley, MA Rouse, A Bateman, JB Rowe

## Abstract

**State of the Art:** Apathy and impulsivity are common in syndromes associated with frontotemporal lobar degeneration (FTLD). They are associated with high carer distress and poor patient outcomes. There are limited treatment options and progress has been hindered by a lack of appropriate outcome measures. This study aimed to develop a carer-rated questionnaire oriented to people with syndromes associated with FTLD.

**Methodology:** Principal component and Rasch analysis were conducted on carer-, clinician- and patient-reported questionnaires and performance-based tests of behavioural change in the “Pick’s Disease and Progressive Supranuclear Palsy Prevalence and Incidence” (PiPPIN) study. We identified two key components which informed subsequent item development for a novel scale which we call the Cambridge Questionnaire for Apathy and Impulsivity Traits (CamQUAIT). The resulting scale comprised two subscales assessing “motivation and support” (CamQUAIT-M) and “impulsivity and challenging behaviours” (CamQUAIT-C). An independent sample of 132 carers for people with FTLD-associated syndromes completed the CamQUAIT, along with a battery of existing measures. The CamQUAIT was reduced to 15 items following Rasch analysis.

**Results:** Both subscales showed good construct validity as assessed by high Person separation index (CamQUAIT-M=0.9; CamQUAIT-C=0.7) and Cronbach’s alpha (CamQUAIT-M=0.9; CamQUAIT-C=0.8). The subscales correlated moderately with each other (r=0.376, p<0.001), and with existing measures of behavioural change.

**Conclusion:** The CamQUAIT is a targeted measurement tool to assess apathy, impulsivity, and related behavioural change in the context of FTLD-related syndromes. The scale demonstrates good measurement properties and is sensitive to group differences, providing a suitable outcome measure to evaluate novel symptomatic treatments.

## Introduction

Apathy is common in neurological and psychiatric diseases (Husain & Roiser, 2018) and consistently linked to poor prognosis (Lansdall et al., 2019; Murley et al., 2021), high caregiver burden (Merrilees et al., 2013), faster cognitive decline (Dujardin et al., 2009), and reduced quality of life (Hollocks et al., 2015). Impulsivity is equally common, and can manifest as risky decisions, falls, excessive gambling, hyper-sexuality, inappropriate social conduct, and binge eating. Impulsive behaviours are difficult to manage, cause significant carer distress (Leroi et al., 2012) and are associated with a poor prognosis (Lansdall et al., 2019; Murley et al., 2021). Apathy and impulsivity are especially prevalent features of syndromes associated with frontotemporal lobar degeneration (FTLD) and frequently coexist (Kok et al., 2021; Lansdall et al., 2017). Syndromes of FTLD include a variety of neurological conditions, here referencing behavioural variant frontotemporal dementia, primary progressive aphasias, progressive supranuclear palsy, and corticobasal syndrome.

Despite increased awareness of the negative impact of these complex behavioural changes, they remain under recognised and poorly treated (Azhar et al., 2022; Nardell & Tampi, 2014; Theleritis et al., 2018). There are a number of potential reasons for this, including challenges associated with current clinical rating scales.

First, there is a lack of appropriately targeted, disease-specific assessment tools to quantify apathy and impulsivity within the FTLD population. Available assessment tools for apathy and impulsivity are widely-used and well-established in other disease groups (Bland et al., 2016; Clarke et al., 2011; Dalley et al., 2011), but may not be appropriate for people affected by FTLD-syndromes. A disease-specific measure should be developed and validated within the intended target population, capturing concepts (symptoms and impacts) that are considered relevant and meaningful to those living with the disease (FDA, 2024). Such measures are rare in FTLD-associated syndromes, limiting meaningful engagement with patients and hindering both assessment and treatment of these debilitating symptoms. For example, in the Barratt Impulsivity Scale (Patton et al., 1995), work-related questions such as “I change jobs” or “I plan for job security” are arguably irrelevant in this disease context as many patients are retired or otherwise unable to work. Other items, such as “I squirm at plays or lectures” contain low frequency words (“squirm”) which are difficult for some FTLD patients to understand (Jefferies & Lambon Ralph, 2006; Lambon Ralph et al., 1998). Assessment tools should be accessible to the patient population and, given the increased recognition of heterogeneity within and between FTLD-associated syndromes (Murley et al., 2020), the use of a transdiagnostic scale to capture symptom commonalities across disease groups could be particularly beneficial for targeted assessment tools.

Second, apathy and impulsivity are often assessed in isolation, despite evidence for their coexistence. People with FTLD-associated syndromes can be both apathetic and impulsive (Kok et al., 2021; Lansdall et al., 2017; Passamonti et al., 2018), suggesting overlapping domains within these multifaceted behavioural constructs. Traditional dopaminergic theories which place these constructs at opposite ends of a motivational spectrum have recently been questioned in conditions associated with FTLD, due to the positive correlation between apathy and impulsivity and the limited clinical efficacy of dopaminergic medication in treating apathy (Azhar et al., 2022). Dopamine deficits and apathy are related but not synonymous (Le Heron et al., 2019), nor incompatible with impulsivity. Measurement tools for these symptoms would therefore benefit from sensitivity to both apathy and impulsivity.

Finally, patient-reported measures may be unreliable in FTLD populations due to characteristic impairments in cognition (Rascovsky et al., 2011) that in many cases lead to lack of insight, likely contributing to discrepancies observed between patient- and caregiver-reported assessments (Lansdall et al., 2017; Massimo et al., 2013). It is particularly challenging in progressive neurodegenerative conditions to determine the point at which patient insight is lost. People with FTLD-associated disorders may also engage in different ways with self-report leading to responses that can be characterised as ‘careless’ (Curran, 2016) or internally inconsistent (Williams et al., 2023). While the patient perspective is critical in identifying important and/or bothersome symptoms and impacts of their disease, caregiver-reported measures may more reliably capture change in these over time. Assessment of these behaviours and their underlying neural correlates may therefore be more appropriately captured by carer-rather than patient-reported measures in the symptomatic stages of disease (Lansdall et al., 2017, 2018).

In this study, we aim to address these challenges by developing a novel, FTLD-specific, carer-rated questionnaire for apathy, impulsivity, and related behavioural change: the Cambridge Questionnaire for Apathy and Impulsivity Traits (CamQUAIT). We aim to provide a valid and reliable assessment tool capturing concepts that are relevant and meaningful to people living with FTLD. We employed Rasch analysis, and psychometric modelling, to empirically test measurement properties of this novel assessment tool within the context of FTLD.

## Methods

### Cohort

The CamQUAIT item set was informed by data from the Pick’s Disease and Progressive Supranuclear Palsy Prevalence and Incidence study (PiPPIN; Coyle-Gilchrist et al., 2016). The PiPPIN study is an epidemiological study of FTLD syndromes in Cambridgeshire and Norfolk with data from 115 patients with FTLD-associated syndromes being used to inform item set development. Patients were diagnosed according to consensus criteria (Armstrong et al., 2013; Gorno-Tempini et al., 2011; Höglinger et al., 2017; Rascovsky et al., 2011).

Once the initial item set was derived, 132 carers of people with FTLD-associated disorders completed a paper and pencil version of the CamQUAIT at the Cambridge Centre for Frontotemporal Dementia. Patient participants provided written informed consent if they had mental capacity to do so or participated following consultation with a personal consultee in accordance with UK law.

### Initial Item Development

A data-driven approach was used for initial item development. In our previous papers, we report the results of a principal component analysis (PCA) on twenty-two questionnaires and performance-based measures assessing apathy, impulsivity, and related behavioural change, gaining insight from multiple perspectives including patient, carer, clinician, and objective tasks (Lansdall et al., 2017). The PCA revealed that carer-rated items relating to apathy, everyday skills and self-care, and carer-rated items relating to challenging behaviours (as measured by the Cambridge Behavioural Inventory-Revised and the Apathy Evaluation Scale) loaded onto distinct components and had dissociable underlying neural correlates (Lansdall et al., 2017; Passamonti et al., 2018). We considered all 63 items loading onto these carer-rated components. After review and removal of redundant and/or repetitive items, 35 items remained for further analysis.

PCA was carried out in SPSSv22 on the remaining 35 items using data from 115 patients. The correlation matrix was used for component extraction, followed by Kaiser-Meyer-Olkin and Bartlett’s test of sphericity to determine the adequacy of the sample for PCA. Varimax rotation ensured orthogonality and maximised dispersion of loadings within components to facilitate interpretation. Selection of components was based primarily on Cattel’s Criteria (Cattell, 1966), extracting components to the left of inflexion on the scree plot. Kaiser’s Criterion (Eigenvalues > 1) was also considered but led to the inclusion of an increased number of weaker components that only explained a small percentage of the variance. For the purposes of scale development, emphasis was placed on the major components which accounted for the majority of the variance.

The principal components analysis revealed three components accounting for >50% of the variance, relating to 1) motivation and support (daily activities, motivation, interests), 2) challenging behaviours (irritability, aggression, impulsivity, and embarrassing behaviours) and 3) interactions with friends.

### Assessment Battery

Carers were invited to complete the CamQUAIT, the Cambridge Behavioural Inventory-Revised (CBI-R; Wear et al., 2008) and the Apathy Evaluation Scale (AES; Marin et al., 1991). The CBI-R is an informant-rated scale designed to assess behavioural change across a range of disorders, while the AES focusses on assessing the behavioural, cognitive, and emotional domains of apathy.

### Rasch Analysis and Benchmarks

Rasch analysis was carried out in RUMM2030. In keeping with the standard benchmarks for acceptable measurement properties, appropriate fit to the Rasch Model was based on several criteria:

1. Item and person fit (within ±2.5 fit residuals, non-significant χ^2^ statistic)
2. Non-significant item-trait interaction (non-significant overall χ^2^ statistic)
3. The t-test protocol for multidimensionality (<5% significant tests and/or lower 95% CI intervals <0.05 when analysing the first residual after removing the “Rasch” component)

Construct validity was assessed using the Person Separation Index and Cronbach’s Alpha. Convergent validity was assessed by exploring the correlation between the CamQUAIT and existing measures of behavioural change and disease severity as listed above.

Before removing items based on Rasch analysis, their theoretical/clinical importance was also considered. Emphasis was placed on removing highly misfitting items rather than rescoring, therefore retaining a consistent scoring structure. Given the origin of the CamQUAIT in a PCA, we anticipated that components would be approaching orthogonality and that the generated subscores would therefore be largely independent. However, given the generation of new items and the inclusion of two “friends” items from a smaller third component, we set a benchmark of <20% shared variance between the subscores as an acceptable degree of dissociation.

Rasch analysis was run independently on the first two components identified in the initial PCA: motivation and support, and challenging behaviours. Only three items contributed strongly to the weak third component, and therefore were not subject to Rasch analysis as this is not recommended for item sets smaller than ten (Lindsay et al., 1991).

This process was followed by structured design of 22 newly derived items encompassing the two main domains identified in the Rasch analysis. The first subscore “motivation and support” (CamQUAIT-M) included 13 items relating to motivation, empathy, and interaction with friends. The second subscore “challenging behaviours” (CamQUAIT-C) contained 9 items relating to impulsivity, aggression, and cooperation. Positive and negative syntax were used in the design of new items. Items were scored on a 4-point Likert scale ranging through Never [0], Sometimes [1], Often [2] and Always [3]. High scores indicate increased levels of behavioural change. Carers were asked to respond based on behaviours seen during “recent weeks”.

## Results

### Cohort

Demographic and clinical characteristics of the development cohort are presented in Table 1 alongside details of 11 healthy controls used in analysis.

**Table 1.** demographics and clinical characteristics of the CamQUAIT cohort. Abbreviations are as follows: PSP Progressive Supranuclear Palsy, CBS Corticobasal Syndrome, PPA Primary Progressive Aphasia, BVFTD Behavioural Variant Frontotemporal Dementia, MND Motor Neurone Disease, CBI-R Cambridge Behavioural Inventory Revised, AES Apathy Evaluation Scale.

| Diagnosis | PSP | CBS | PPA | BVFTD<br>(±MND) | MND | Controls |
| --- | --- | --- | --- | --- | --- | --- |
| **N** | 54 | 17 | 20 | 24 | 17 | 11 |
| **Age** | 72.13 ± 6.90 | 72.47 ± 8.25 | 70.80 ± 5.94 | 61.07 ± 12.58 | 66.46 ± 9.66 | 66.09 ± 5.54 |
| **Gender**<br>(M:F) | 31:23 | 4:13 | 9:11 | 16:8 | 12:5 |  |
| **CBI-R** | 64.48 ± 35.86 | 59.38 ± 34.14 | 74.06 ± 35.64 | 103.43 ± 27.62 | 18.59 ± 25.15 | - |
| **AES** | 48.23 ± 12.22 | 39.46 ± 15.12 | 48.55 ± 13.92 | 57.69 ± 9.77 | - | - |

### Rasch Analysis

Independent Rasch analyses were conducted on each subscore and their item set. Initial development using PCA provided strong support that the generated subscores related to independent behavioural domains and including all items in a single Rasch analysis revealed strong multidimensionality.

#### Missing Data

Of the 132 CamQUAIT datasets entered into the Rasch analysis, 13/132 (9.8%) had unmarked or missing data for CamQUAIT-M and 20/132 (15%) for CamQUAIT-C. The CamQUAIT-M had two items with no missing data. The remaining items had less than five missing data points, with the exception of item 5 “likes to get things done during the day” (N=7) and item 7 “has an intense approach to life” (N=8). CamQUAIT-C had four items with no missing data, while the remaining had less than or equal to 5 missing data points. The most frequently unmarked item was item 18 “is unenthusiastic about his/her usual hobbies”.

#### Construct Validity

##### CamQUAIT-M: Motivation and Support

A Person Separation Index and Cronbach’s alpha of 0.9 indicated good separation of items along the construct and sufficient power to discriminate between 4 groups of respondents (Fischer, 1992).

The scale showed good overall fit to the Rasch model in terms of item-person interaction statistics (item mean = 0.297, SD = 1.898; person mean = -0.155, SD = 1.274), although the item-trait interaction was significant (χ^2^= 96.625, p<0.001) indicating deviation from the model expectations. Item 7 “has an intense approach to life” and item 13 “spends his/her day doing things of interest to him/her” were removed due to high fit statistics of 3.03 and 4.36 respectively (>±2.5), and highly significant Chi squared values (p<0.001). Item 20 “is interested in having friends” showed differential item functioning by gender and was removed following an unsuccessful attempt to split the item (which can often result in the item fitting the overall item set). Item 11 “is interested in doing new things” demonstrated high residual correlation with item 19 “likes to learn new things”. After examining improvement in fit following the removal of each item in turn, item 11 was removed.

This resulted in an overall fit to the Rasch model, including good item-person fit statistics (item mean = 0.072, SD = 1.269; person mean = -0.200, SD = 1.093), a non-significant item-trait interaction (χ^2^ = 27.5, df = 18, p>0.05), and a high Person Separation Index of 0.9 (see Table 2).

**Table 2.** Rasch model statistics for the two subscores of the CamQUAIT: the motivation and support subscore (CamQUAIT-M) and the challenging behaviours subscore (CamQUAIT-C). *N omitting test-based extreme person estimates.

| Analysis | | Items | Psi/$\alpha$ | Item fit residual | | Person fit residual | | Chi square interaction | | Unidimensionality t-tests (ci) | | |
| --- | --- | --- | --- | --- | --- | --- | --- | --- | --- | --- | --- | --- |
|  |  |  |  | Mean | SD | Mean | SD | Value (df) | P | Perc<5% | N* | Lower CI |
| CamQUAIT-M | Initial | 13 | 0.9/0.9 | 0.298 | 1.898 | 0.155 | 1.274 | 96.625 (26) | <0.0001 | 14 | 130 | 0.70 |
|  | Final | 9 | 0.9/0.9 | 0.072 | 1.269 | 0.200 | 1.093 | 27.508 (18) | 0.070 | 9 | 128 | 0.33 |
| CamQUAIT-C | Initial | 9 | 0.7/0.8 | 0.100 | 1.890 | 0.164 | 0.948 | 75.460 (18) | <0.0001 | 8 | 131 | 0.024 |
|  | Final | 6 | 0.7/0.8 | 0.027 | 0.992 | 0.222 | 0.872 | 10.634 (12) | 0.560 | 7 | 119 | 0.020 |

Unidimensionality of the reduced item set was assessed using principal components analysis of the person residuals. The first residual accounted for 21% of the variance, once the “Rasch” factor was removed. The item set showed acceptable unidimensionality (Tennant & Conaghan, 2007), with significant differences between the two item subsets at the 5% level for 9/128 (7%) persons with a lower 95% confidence interval proportion of 0.033 (<0.05), omitting test-based extremes.

##### CamQUAIT-C: Challenging Behaviours

A Person Separation Index of 0.7 and Chronbach’s alpha of 0.8 indicated good ability to statistically differentiate between 2-3 groups.

The scale showed acceptable person fit statistics (mean = -0.164, SD = 0.950) but high item standard deviation (mean = 0.100, SD = 1.890) and a significant item-trait interaction (χ^2^= 75.460, df = 18, p<0.001). No items displayed significant differential item functioning by gender. Two items showed strong misfit with high fit residuals and significant χ^2^ values and were removed – i.e. item 18 “is unenthusiastic about his/her usual hobbies” (Fit Res = 3.22, χ^2^ p<0.05) and item 16 “is tearful or cries” (Fit Res = 2.94, χ^2^ p<0.001). Item 14 “has a poor understanding of his/her problems” was removed after theoretical examination and consideration of its disordered response structure, resulting in a significant improvement in fit (item mean = 0.027, SD = 0.992; person mean = -0.222, SD=0.872) and a non-significant χ^2^ of 10.6 (p=560). Note that rescoring item 14 did not significantly improve fit.

The reduced item set successfully met the t-test protocol for unidimensionality, with the first residual accounting for 35% of the variance and significant differences between the two item subsets at the 5% level for 7/119 (5.9%) persons with a lower 95% confidence interval proportion of 0.020 (<0.05), omitting test-based extremes (see Table 2).

### Scale Reconstruction and Scoring

The final item set consisted of 15 items, 9 pertaining to the motivation and support subscore (CamQUAIT-M) and 6 to the challenging behaviours subscore (CamQUAIT-C). The final item set and accompanying scoring sheet can be found in the Supplementary Material.

Item scoring was retained with 4 categories: Never [0], Sometimes [1], Often [2] and Always [3]. Although some items showed some disordered thresholds, a larger sample size is required to ensure efficient utilisation of all response categories. Maintaining scoring structure facilitates further data collection and the consolidation of all datasets.

Table 3 includes the score to logit to severity conversions. The Person Separation Index and Cronbach’s alpha of CamQUAIT-M indicated ability to dissociate between 4 groups, but for consistency, severity levels for both subscores were split into 3 categories based on the lower Person Separation Index and Cronbach’s alpha of CamQUAIT-C. Severity categories ranged from Mild to Moderate to Severe, with 9 levels per category for CamQUAIT-M and 6 levels for CamQUAIT-C.

**Table 3.** CamQUAIT conversion between section scores, logit and proposed severity rating.

| Motivation and Support |  |  | Challenging Behaviours |  |  |
| --- | --- | --- | --- | --- | --- |
| *Score* | *Logit* | *Category* | *Score* | *Logit* | *Category* |
| 0 | -3.842 | Normal | 0 | -4.039 | Normal |
| 1 | -3.079 | Normal | 1 | -3.102 | Normal |
| 2 | -2.538 | Normal | 2 | -2.408 | Normal |
| 3 | -2.154 | Normal | 3 | -1.894 | Normal |
| 4 | -1.848 | Normal | 4 | -1.472 | Normal |
| 5 | -1.591 | Normal | 5 | -1.107 | Normal |
| 6 | -1.367 | Normal | 6 | -0.779 | Normal |
| 7 | -1.168 | Normal | 7 | -0.477 | Intermediate |
| 8 | -0.987 | Normal | 8 | -0.192 | Intermediate |
| 9 | -0.82 | Normal | 9 | 0.082 | Intermediate |
| 10 | -0.662 | Intermediate | 10 | 0.351 | Intermediate |
| 11 | -0.511 | Intermediate | 11 | 0.62 | Intermediate |
| 12 | -0.363 | Intermediate | 12 | 0.893 | Intermediate |
| 13 | -0.217 | Intermediate | 13 | 1.178 | Severe |
| 14 | -0.071 | Intermediate | 14 | 1.488 | Severe |
| 15 | 0.08 | Intermediate | 15 | 1.84 | Severe |
| 16 | 0.239 | Intermediate | 16 | 2.273 | Severe |
| 17 | 0.407 | Intermediate | 17 | 2.878 | Severe |
| 18 | 0.589 | Intermediate | 18 | 3.723 | Severe |
| 19 | 0.791 | Severe |  |  |  |
| 20 | 1.018 | Severe |  |  |  |
| 21 | 1.276 | Severe |  |  |  |
| 22 | 1.574 | Severe |  |  |  |
| 23 | 1.924 | Severe |  |  |  |
| 24 | 2.348 | Severe |  |  |  |
| 25 | 2.895 | Severe |  |  |  |
| 26 | 3.686 | Severe |  |  |  |
| 27 | 4.829 | Severe |  |  |  |

CamQUAIT /27 M
CamQUAIT /18 C

The CamQUAIT-M subscale (items 1, 3, 5, 8, 11-15) is reversed for valence. For ease of scoring the subscale of each question is also indicated on the questionnaire.

### CamQUAIT Performance by Group and Disease Severity

Significant differences in performance on the CamQUAIT subscores were observed across diagnostic groups, with the motor neuron disease cohort scoring lowest on average and the bvFTD cohort receiving the highest scores (see Figure 1).

**Figure 1.**
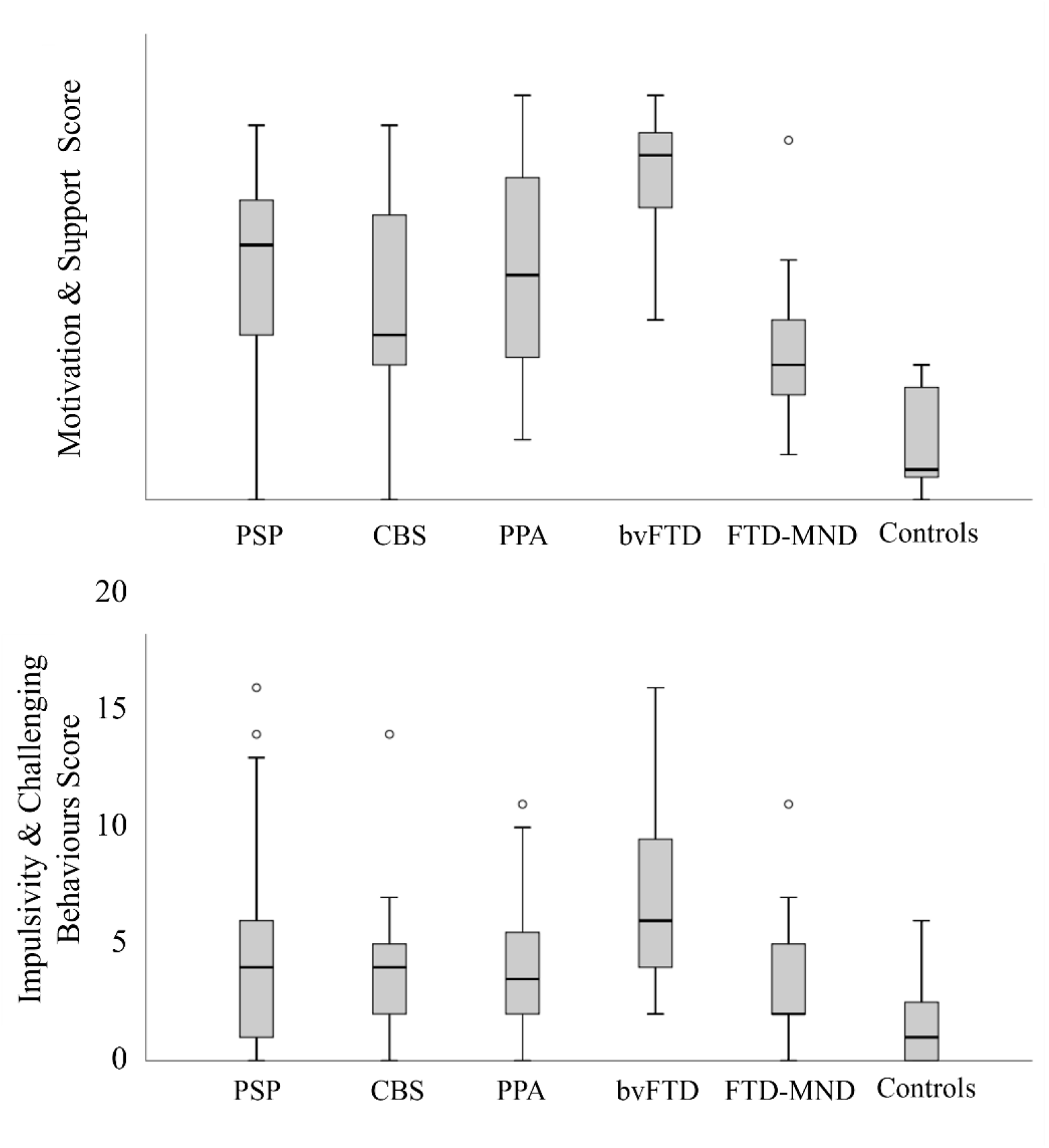
Box plot of scores from the two subscales of the CamQUAIT across disease group. (A) Shows scores on the motivation and support subscale (CamQUAIT-M), while (B) shows scores on the impulsivity and challenging behaviours subscore (CamQUAIT-C). PSP = Progressive Supranuclear Palsy CBS = Corticobasal Syndrome PPA = Primary progressive Aphasia bvFTD = behavioural variant Frontotemporal Dementia FTD-MND = Frontotemporal Dementia with Motor Neuron Disease

The CamQUAIT subscores “motivation and support” (CamQUAIT-M) and “challenging behaviours” (CamQUAIT-C) were positively correlated across groups (r=0.376, p<0.001) but met our initial criteria of <20% shared variance (14% shared variance).

There was a positive relationship between the CamQUAIT and other widely used measures of behavioural change, such as the Revised Cambridge Behavioural Inventory (CamQUAIT-M: r= 0.663, p<0.001; CamQUAIT-C: r=0.595, p<0.001) and the Apathy Evaluation Scale (CamQUAIT-M: r= 0.909, p<0.001; CamQUAIT-C: r=0.376, p<0.001). This correlation was stronger when comparing the CamQUAIT-M with measures of motivation and apathy than challenging behaviours, and vice versa for the CamQUAIT-C.

## DISCUSSION

The Cambridge Questionnaire for Apathy and Impulsivity Traits (CamQUAIT) is a novel, targeted measurement tool to assess apathy, impulsivity, and related behavioural change in the context of FTLD-related disorders. The scale demonstrates good measurement properties, meeting the assumptions of unidimensionality under the Rasch model and showing good overall sensitivity to change in apathy and impulsivity across the FTLD spectrum. The CamQUAIT may therefore aid in overcoming current hurdles to assessing apathy and impulsivity within this disease group.

The CamQUAIT has several potential advantages over existing measures. It is a short and simple assessment tool which demonstrates empirically tested psychometric validity for the assessment of apathy and impulsivity in the context of FTLD-related disorders. Most carers were able to complete the CamQUAIT easily, and the most frequently unmarked item from each subscore was removed during the Rasch processing steps. Overall, items had a low number of missing data, ranging from 0-5 unmarked items across the 132 respondents (<4%). The scale demonstrated good construct validity, correlating with other widely used measures of behavioural change such as the Cambridge Behavioural Inventory and Apathy Evaluation Scale.

In contrast to some alternative assessment tools which are widely used - but developed principally for psychiatric or healthy populations - the CamQUAIT contains items that are relevant at face value to people with advancing cognitive and motor disability. The logit scoring system provides a measurement scaling system for assessment, examining the trade-off between respondent “ability” (behaviour, personality traits, etc.) and item difficulty, placing items and people along the same continuum by converting raw data into equal interval logit scores. This approach has gained traction in some groups, scoring individuals based on their ability level, while items are scored based on their difficulty, according to performance of the whole sample. By meeting the assumptions of the Rasch model, one can be confident that the items in a given questionnaire assess the same latent trait within the chosen population.

The CamQUAIT item set was derived from data collected from FTLD patients’ informants only, making it a targeted assessment. Carers of people with bvFTD consistently reported the highest scores while carers of people with motor neuron disease report the lowest. CamQUAIT-M revealed strong endorsement of motivational deficits relating to everyday activities and interactions with friends across diagnostic groups, in line with our previous studies (Lansdall et al., 2017). In contrast, the CamQUAIT-C revealed predominance of challenging behaviours in bvFTD and PSP groups, consistent with our previous findings and the broader literature emphasising the frequency of such behaviours in bvFTD (Hughes et al., 2011; O’Callaghan et al., 2013; Rascovsky et al., 2011) and PSP (O’Sullivan et al., 2010; Rittman et al., 2016; Zhang et al., 2016), for which they also form part of the diagnostic criteria. Indeed, the latest diagnostic criteria for PSP recognised impulsivity as a core feature, acknowledging PSP-frontal variant (PSP-F) which is characterised by predominant behavioural change (Höglinger et al., 2017). The scale also effectively captured the full spectrum of severity from normal to severe, indicated by good item-person fit of the scale subscores. Moreover, the scale remains applicable transdiagnostically as all groups scored higher on average than controls, indicating endorsement of these behaviours across groups to varying extents.

Finally, although the CamQUAIT-M and CamQUAIT-C subscores were positively correlated, the correlation was sufficiently low (<15% shared variance) to justify two subscores and supported by high multidimensionality when including all items in a single Rasch analysis.

This study has several limitations. A rescoring process was not repeated to identify and resolve residual misfit. In view of the relatively small sample size, particularly at a diagnostic group level, it was considered that there was insufficient data to warrant item further rescoring due to the risk of categories working ineffectively. A larger sample may result in appropriate endorsement of all response categories, and we therefore prioritised the maintenance of a simple and consistent scoring structure. In addition, not all psychometric properties were evaluated in this study, nor was direct input from patients and caregivers collected in item development, for example on ‘meaningfulness’ or ‘impact’ of items and their underlying behaviours. The latter could help to confirm the meaningfulness of the CamQUAIT for those living with FTLD-related syndromes and be a useful future step.

In summary, studies assessing the benefit of symptomatic treatments for apathy and impulsivity in FTLD-associated syndromes should use appropriate tools with established measurement properties for the population in question. In this paper, we demonstrate that the novel 9-item and 6-item CamQUAIT subscores meet the Rasch model expectations and demonstrate good validity for the assessment of apathy and impulsivity in these syndromes. We hope that the CamQUAIT will be a useful addition for studies evaluating the benefit of potential new symptomatic treatments for apathy and impulsivity in FTLD-associated disorders.

## Data Availability

For enquiries related to source data, please contact the corresponding author. A material transfer agreement may be required, to comply with the terms of consent and GDPR.

## Acknowledgements

This work has been funded by the Medical Research Council (MC_UU_00030/14; MR/T033371/1), the Wellcome Trust (220258), the NIHR Cambridge Biomedical Research Centre (NIHR203312), the Cambridge Centre for Parkinson-plus and the Holt fellowship. The views expressed are those of the authors and not necessarily those of the NIHR or the Department of Health and Social Care. For the purpose of open access, the authors have applied a CC BY public copyright licence to any Author Accepted Manuscript version arising from this submission.

