## Supplemental Material for "The Cambridge Questionnaire for Apathy and Impulsivity Traits (CamQUAIT): a novel assessment tool for frontotemporal lobar degeneration-related syndromes"

Your Name: ............................................................ Date: ..............................

Patient's Name: ............................................................

Relationship to Patient: ............................................................

Please answer the following questions about behaviour you have seen in recent weeks.

Circle the word that best applies (never, sometimes, often, always) for every question.

| **The patient named above:** | **Never** | **Sometimes** | **Often** | **Always** | M | **C** |
|---|---|---|---|---|---|---|
| 1. feels it is important to finish a job/task | Never | Sometimes | Often | Always | | |
| 2. is uncooperative | Never | Sometimes | Often | Always | | |
| 3. shows initiative | Never | Sometimes | Often | Always | | |
| 4. has outbursts of aggression (or temper) | Never | Sometimes | Often | Always | | |
| 5. likes to get things done during the day | Never | Sometimes | Often | Always | | |
| 6. is socially embarrassing | Never | Sometimes | Often | Always | | |
| 7. is impulsive (acts without thinking) | Never | Sometimes | Often | Always | | |
| 8. shows interest in things | Never | Sometimes | Often | Always | | |
| 9. is easily irritated | Never | Sometimes | Often | Always | | |
| 10. makes insensitive or tactless comments | Never | Sometimes | Often | Always | | |
| 11. is considerate of other family members and their concerns | Never | Sometimes | Often | Always | | |
| 12. can manage each day without being told what to do | Never | Sometimes | Often | Always | | |
| 13. likes to learn new things | Never | Sometimes | Often | Always | | |
| 14. gets excited when something good happens to him/her | Never | Sometimes | Often | Always | | |
| 15. is interested in meeting friends | Never | Sometimes | Often | Always | | |

**SCORING**

All items marked as part of the ‘Motivation and Support’ subscale are reverse scored. Subscores are indicated by the shaded boxes to the right of the scale.

Items are scored as follows*:

0 = Never

1 = Sometimes

2 = Often

3 = Always

*For reverse scored items, 3 = Never, 2 = Sometimes, 1 = Often, 0 = Always.

Subscores:

- Motivation and Support Sub-score (9 items):....../27
- Impulsivity and Challenging Behaviours Sub-score (6 items):...../18
- Total score (for clinical purposes only):...../45

Higher scores indicate greater impairment, reflecting increased behavioural change including apathy/impulsivity.
